# Dasatinib nephrotoxicity correlates with patient-specific pharmacokinetics

**DOI:** 10.1101/2023.04.09.23288333

**Authors:** Benjamin O. Adegbite, Matthew H. Abramson, Victoria Gutgarts, Marcel F. Musteata, Kinsuk Chauhan, Alecia N. Muwonge, Kristin A. Meliambro, Steven P. Salvatore, Sebastian El Ghaity-Beckley, Marina Kremyanskaya, Bridget Marcellino, John O. Mascarenhas, Kirk N. Campbell, Lili Chan, Steven G. Coca, Ellin M. Berman, Edgar A. Jaimes, Evren U. Azeloglu

**Affiliations:** Division of Nephrology, Department of Medicine, Icahn School of Medicine at Mount Sinai, New York, NY; Internal Medicine, Mount Sinai Morningside/West, New York, NY; Renal Service, Memorial Sloan Kettering Cancer Center, New York, NY; Department of Pharmaceutical Sciences, Albany College of Pharmacy & Health Sciences, Albany, NY; Clinical Pathology and Laboratory Medicine, Weill Cornell Medical College, New York, NY; Leukemia Service, Memorial Sloan Kettering Cancer Center, New York, NY; Division of Hematology/Medical Oncology, Tisch Cancer Institute, Icahn School of Medicine at Mount Sinai, New York, NY; Department of Pharmacological Sciences, Icahn School of Medicine at Mount Sinai, New York, NY

**Keywords:** chronic myeloid leukemia, CML, podocyte, glomerulus, kidney injury

## Abstract

**Introduction:** Dasatinib has been associated with nephrotoxicity. We sought to examine the incidence of proteinuria on dasatinib and determine potential risk factors that may increase dasatinib-associated glomerular injury.

**Methods:** We examine glomerular injury via urine albumin-to-creatinine ratio (UACR) in 101 chronic myelogenous leukemia patients who were on tyrosine-kinase inhibitor (TKI) therapy for at least 90 days. We assay plasma dasatinib pharmacokinetics using tandem mass spectroscopy, and further describe a case study of a patient who experienced nephrotic-range proteinuria while on dasatinib.

**Results:** Patients treated with dasatinib (n= 32) had significantly higher UACR levels (median 28.0 mg/g, IQR 11.5 – 119.5) than patients treated with other TKIs (n=50; median 15.0 mg/g, IQR 8.0 – 35.0; p < 0.001). In total, 10% of dasatinib users exhibited severely increased albuminuria (UACR > 300 mg/g) versus zero in other TKIs. Average steady state concentrations of dasatinib were positively correlated with UACR (ρ = 0.54, p = 0.03) as well as duration of treatment (*p* =0.003). There were no associations with elevated blood pressure or other confounding factors. In the case study, kidney biopsy revealed global glomerular damage with diffuse foot process effacement that recovered upon termination of dasatinib treatment.

**Conclusions:** Exposure to dasatinib is associated a significant chance of developing proteinuria compared to other similar TKIs. Dasatinib plasma concentration significantly correlates with increased risk of developing proteinuria while receiving dasatinib. Screening for renal dysfunction and proteinuria is strongly advised for all dasatinib patients.

## Introduction

Dasatinib is a second generation BCR-ABL1 tyrosine-kinase inhibitor (TKI) that has been used to treat chronic myelogenous leukemia (CML) and acute lymphoblastic leukemia (ALL) harboring the Philadelphia chromosome, a balanced translocation between chromosome 9 and 22, t(9;22). Since the Food and Drug Administration (FDA) approval in 2006, dasatinib has been extensively used as a first- and second-line agent for the treatment of CML ^1,2^.

Dasatinib is associated with adverse drug reactions (ADRs) including headaches, nausea, and rhabdomyolysis ^3^. Nephrotoxicity (in the form of proteinuria) of dasatinib is another ADR that has been reported in case reports, and in the package insert, it is noted that less than 1% of users will experience it as a treatment emergent adverse event (AE) ^4^. As many as 32% of FDA approved drugs have shown post-market AEs; as such, providers may not rigorously monitor the kidney function, especially in the absence of prior kidney disease ^5^. This further becomes important in the setting of kidney injury since continuous insult can eventually progress into chronic kidney disease, kidney failure, and other non-renal outcomes, such as ischemic heart disease and toxin accumulation ^6^. Cancer patients already have an elevated risk for kidney disease; therefore, introducing dasatinib therapy may potentially hasten the process ^7^.

We have previously identified dasatinib as the TKI with one of the highest reported risks of nephrotoxicity ^8^ in the absence of concurrent hypertension through meta-analysis of the FDA AE reporting system ^9^. This implication that dasatinib may have a direct effect on glomerular injury (rather than secondary to hypertension) was further supported by the fact that only dasatinib (and none of the other comparable TKIs) disrupted the actin cytoskeleton and focal adhesion architecture of cultured mouse podocytes *in vitro*. These findings were also recapitulated *in vivo* in a murine model of glomerular injury ^8^.

In patients, several case reports have identified individuals with CML who developed proteinuria while receiving dasatinib therapy ^10,11^. In one case, a 40-year-old Japanese man developed heavy proteinuria with urine protein-creatinine (UPC) ratio of 12.2 and hematuria after receiving 100 mg daily of dasatinib for three months; his kidney biopsy showed extensive podocyte effacement ^12^. After switching treatment to nilotinib, however, his UPC reduced to 0.63, and his hematuria disappeared within two weeks. In another instance, a three-year-old female child developed heavy proteinuria with UPC of 17 after receiving 60 mg of dasatinib for 17 months ^13^. Kidney biopsy also showed podocyte effacement as well as global glomerulosclerosis. Similar to the previous case, the proteinuria stopped two months after dasatinib discontinuation.

We sought to systematically investigate the incidence of dasatinib-associated nephrotoxicity in CML patients receiving treatment with TKIs at Memorial Sloan Kettering Cancer Center (MSKCC) and the Tisch Cancer Institute (TCI) at Mount Sinai Hospital. We also sought to identify the clinical and pharmacological factors that may increase the risk of developing nephrotoxicity while being treated with dasatinib.

## Methods

### Study Population

Institutional Review Boards (IRB) at MSKCC and TCI approved protocol and patient consent presented here. A total of 101 patients with CML from MSKCC and TCl were recruited between October 2018 and April 2022. Consent was acquired prior to enrollment. The TKIs included dasatinib, imatinib, bosutinib, ponatinib, and nilotinib. Patients may have received more than one TKI during the course of their cancer treatment, but data from the most recent drug administered was used for this analysis. Only patients that were receiving a given TKI for over 90 days at the time of sample collection were included in analysis.

### Sample Preparation

Urine from the patients were obtained in collection containers on ice and aliquoted into 15 mL conical centrifuge tubes under aseptic conditions. Tubes were then centrifuged at 4,500 RPM at 4°C for 10 minutes. Afterwards, 1 mL of the filtered urine was transferred into cryotubes; these tubes were sent to internal CLIA-certified clinical labs for urinalysis. Excess urine was then stored at -80°C. When samples (urine and plasma) were collected, patients were each at a different point in their TKI course. In addition, samples were collected once for each patient.

In the cohort using dasatinib, 23 participants were available for blood sample collection. This collection also occurred the same day when their urine was obtained. We acquired data on the time of last administered dose of dasatinib, fasting status, and time of their last meal in relation to the time of blood draw. For plasma preparation, blood samples were homogenized in tubes and centrifuged at 200G for 10 minutes. Then, the separated plasma was spun for 1,000G for another 10 minutes for precipitate to settle. Afterwards, 300 μL of plasma was aliquoted into two cryotubes for each sample under aseptic conditions. Cryotubes were stored at -80°C until analysis.

### Assessment of Proteinuria

Patients experiencing glomerular injury were identified based on level of proteinuria, as determined from a spot urine sample with urine albumin:urine creatinine ratio (UACR) in a CLIA certified laboratory. If albumin concentrations were unmeasurable (i.e., < 0.5 mg/dL), we used a value of 0.1 mg/dL. Proteinuria was indicated as having a UACR of at least 30 μg/mg. Furthermore, as per the KDIGO CKD guidelines ^14^, patients with a UACR of at least 30 μg/mg and less than 300 μg/mg were considered to have moderately increased albuminuria, while patients with UACR of at least 300 μg/mg were classified as severely increased albuminuria. Of note, we also identify patients with proteinuria as *nephrotoxic* in all subsequent tables and figures.

### Assessment of Plasma Dasatinib Concentration by Liquid Chromatography with Tandem Mass Spectrometry (LC-MS/MS)

Plasma samples were processed by protein precipitation with acetonitrile (1:2 plasma:acetonitrile by volume) containing deuterated dasatinib (D_8_) as internal standard at a concentration of 10 ng/mL; the vials were vortexed for 10 min at 2,000 rpm, frozen at -20°C for 30 min, and centrifuged at 20,000 rcf for 30 min. Subsequently, 75 μL supernatant were transferred into total recovery vials for autosampler and subjected to analysis by LC-MS/MS. The chromatography system consisted of a Shimadzu Nexera XR liquid chromatograph equipped with a degasser, binary pump, temperature-controlled autosampler (4°C), and temperature-controlled column oven (40°C). The mass spectrometer consisted of an AB Sciex 5500 QTRAP triple quadrupole linear ion trap mass spectrometer with electrospray ionization source. The autosampler was used to inject 3.0 μL sample onto the column. The injection needle was washed before and after sample introduction with 2.0 mL solution 1:1 methanol: water. The ion source was set to positive ionization with an ion spray voltage of 5000 V. Nitrogen was utilized in the ion source as curtain gas and desolvation gas at a temperature of 500°C. Analytical separation was performed on a Poroshell 120 EC-C18 column, 2.1 × 50 mm, containing 2.7-micron particles (Agilent Technologies). The mobile phases A and B consisted of 0.1% formic acid in water and 0.1% formic acid in acetonitrile, respectively, flowing at 400 μL/min. The gradient program started from 90% A which was decreased to 5% A over 5 min and was returned to the initial percentage of 90% A from 5 to 5.5 min, followed by 1.5 min equilibration with 90% A. Nitrogen at medium flow was utilized for collisionally activated dissociation in the second quadrupole. The analyte and internal standard were quantified in MRM mode with the specific transitions 488.4→401.2 (quantitation) and 488.4→232.1 (confirmation) for dasatinib, and 496.4→406.2 for dasatinib-D_8_. The system was operated with Analyst 1.7 for data acquisition and MultiQuant 3.0.2 for automatic chromatogram processing.

### Pharmacokinetic Analysis

All blood samples for pharmacokinetic analysis were collected after the subjects reached steady state levels since all subjects were receiving current treatment for over 90 days. Dasatinib concentrations were determined using a validated LC-MS/MS method; instrument calibration was done with pooled plasma from healthy volunteers not having received dasatinib. Data analysis was performed individually for each subject. Statistical analysis, curve fitting, and parameter estimation using an oral administration model were performed using the PKanalix application from the Monolix suite (version 2019R2, Lixoft software).

### Statistical Analyses

We present the participants baseline characteristics by their drug status (dasatinib versus non-dasatinib) and also within dasatinib drug group by their proteinuria status. The categorical variables are presented as n (%) and were compared by *X*^2^ test or Fisher’s test as appropriate. The continuous variables are presented as mean (SD) and in skewed distribution, as median (interquartile range, IQR). The Student’s *t* test for normally distributed variables and Wilcoxon Mann-Whitney test for skewed variables were used for the comparison between groups. We compared the association of *log2* transformed UACR with duration of drug versus drug categories using linear regression.

## Results

There was a total of 101 patients enrolled in the study: 95 from MSKCC and 6 from TCI. Nine patients had missing UACR, another five had been receiving TKI therapy for less than 90 days, and five had missing drug regimen information. Thus, the following analyses were conducted on the remaining 82 patients. Thirty-two patients were receiving dasatinib, and fifty were receiving another TKI (31 on imatinib, 7 on bosutinib, 10 on nilotinib, and 2 on ponatinib). In terms of group characteristics, dasatinib users were on average 48 ± 13 years old and slightly more were female (53%), while non-dasatinib users were older on average at 59 ± 12 years, half female (50%) (p < 0.05). A complete summary of demographics, drug regimen information, and the associated clinical parameters are listed in Tables 1 and 2.

**Table 1.**
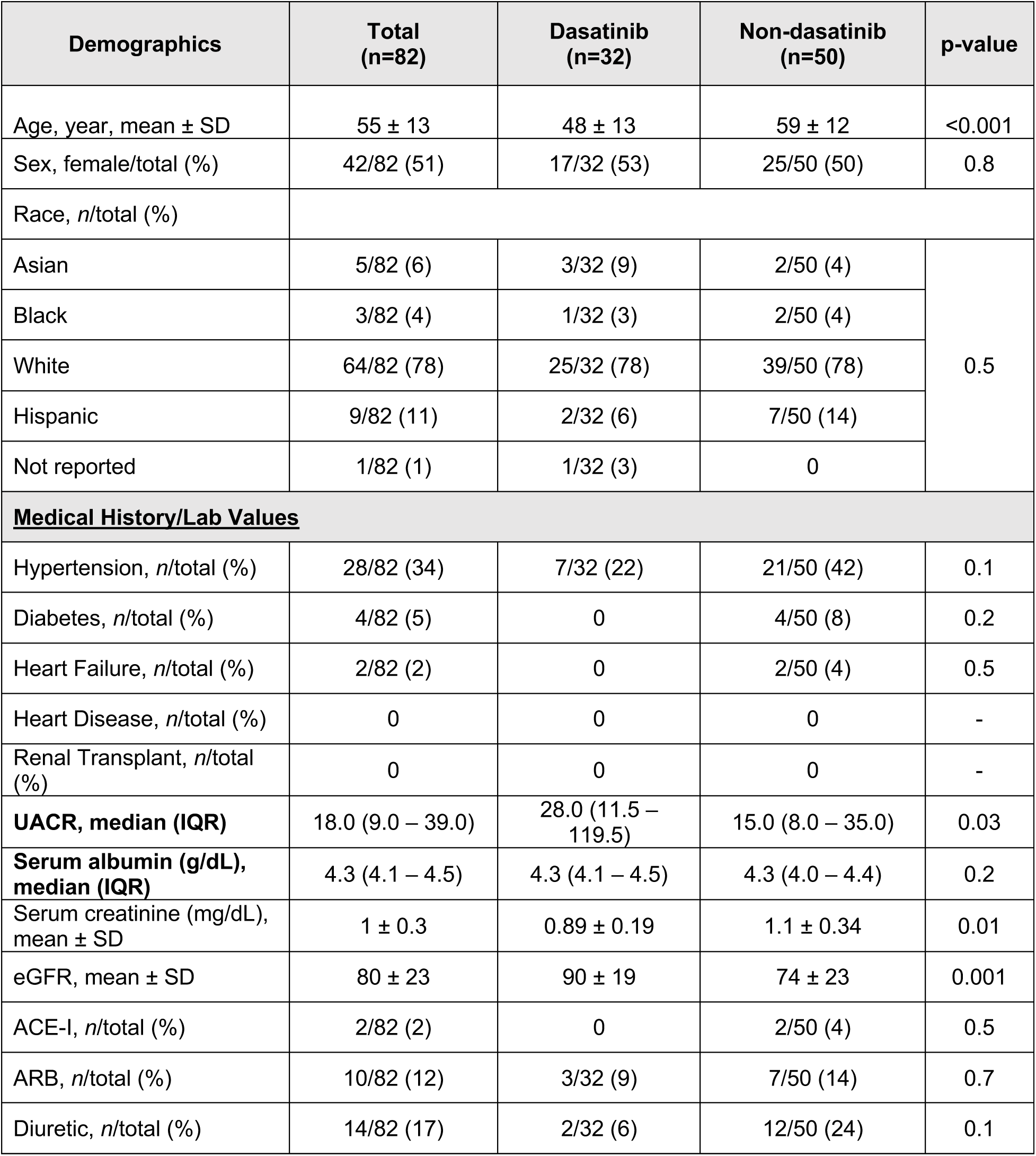
Demographics of patients receiving dasatinib or other TKIs

| Demographics | Total (n=82) | Dasatinib (n=32) | Non-dasatinib (n=50) | p-value |
| --- | --- | --- | --- | --- |
| Age, year, mean ± SD | 55 ± 13 | 48 ± 13 | 59 ± 12 | <0.001 |
| Sex, female/total (%) | 42/82 (51) | 17/32 (53) | 25/50 (50) | 0.8 |
| Race, n/total (%) |  |  |  |  |
| Asian | 5/82 (6) | 3/32 (9) | 2/50 (4) | 0.5 |
| Black | 3/82 (4) | 1/32 (3) | 2/50 (4) |  |
| White | 64/82 (78) | 25/32 (78) | 39/50 (78) |  |
| Hispanic | 9/82 (11) | 2/32 (6) | 7/50 (14) |  |
| Not reported | 1/82 (1) | 1/32 (3) | 0 |  |
| **<u>Medical History/Lab Values</u>** |  |  |  |  |
| Hypertension, n/total (%) | 28/82 (34) | 7/32 (22) | 21/50 (42) | 0.1 |
| Diabetes, n/total (%) | 4/82 (5) | 0 | 4/50 (8) | 0.2 |
| Heart Failure, n/total (%) | 2/82 (2) | 0 | 2/50 (4) | 0.5 |
| Heart Disease, n/total (%) | 0 | 0 | 0 | - |
| Renal Transplant, n/total (%) | 0 | 0 | 0 | - |
| **UACR, median (IQR)** | 18.0 (9.0 – 39.0) | 28.0 (11.5 – 119.5) | 15.0 (8.0 – 35.0) | 0.03 |
| **Serum albumin (g/dL), median (IQR)** | 4.3 (4.1 – 4.5) | 4.3 (4.1 – 4.5) | 4.3 (4.0 – 4.4) | 0.2 |
| Serum creatinine (mg/dL), mean ± SD | 1 ± 0.3 | 0.89 ± 0.19 | 1.1 ± 0.34 | 0.01 |
| eGFR, mean ± SD | 80 ± 23 | 90 ± 19 | 74 ± 23 | 0.001 |
| ACE-I, n/total (%) | 2/82 (2) | 0 | 2/50 (4) | 0.5 |
| ARB, n/total (%) | 10/82 (12) | 3/32 (9) | 7/50 (14) | 0.7 |
| Diuretic, n/total (%) | 14/82 (17) | 2/32 (6) | 12/50 (24) | 0.1 |

**Table 2.** Demographics of nephrotoxic vs non-nephrotoxic dasatinib patients

| Demographics | Total (n=32) | Nephrotoxic (n=8) | Non- Nephrotoxic (n=24) | p-value |
| --- | --- | --- | --- | --- |
| Age, yr, mean ± SD | 48 ± 13 | 50 ± 12 | 48 ± 14 | 0.7 |
| Sex, female/total (%) | 17/32 (54) | 5/8 (63) | 12/24 (50) | 0.7 |
| Race, n/total (%) |  |  |  |  |
| Asian | 3/32 (9) | 0 | 3/24 (13) | 0.2 |
| Black | 1/32 (3) | 1/8 (13) | 0 |  |
| White | 25/32 (78) | 6/8 (75) | 19/24 (80) |  |
| Hispanic | 2/32 (6) | 0 | 2/24 (8) |  |
| Not reported | 1/32 (3) | 1/8 (13) | 0 |  |
| **<u>Medical History/Lab Values</u>** |  |  |  |  |
| Hypertension, n/total (%) | 7/32 (22) | 2/8 (25) | 5/24 (21) | 1.0 |
| Diabetes, n/total (%) | 0 | 0 | 0 | - |
| Heart Failure, n/total (%) | 0 | 0 | 0 | - |
| Heart Disease, n/total (%) | 0 | 0 | 0 | - |
| Renal Transplant, n/total (%) | 0 | 0 | 0 | - |
| UACR, median (IQR) | 28.0 (11.5 – 119.5) | 273.5 (183.0 – 596.0) | 16.5 (8.0 – 37.0) | <0.001 |
| Serum albumin (g/dL), **median (IQR)** | 4.3 (4.1 – 4.5) | 4.5 (4.3 – 4.6) | 4.2 (4.1 – 4.5) | 0.4 |
| Serum creatinine (mg/dL), mean ± SD | 0.89 ± 0.19 | 0.96 ± 0.31 | 0.87 ± 0.12 | 0.2 |
| eGFR, ml/min/1.73 m<sup>2</sup>, mean ± SD | 90 ± 19 | 85 ± 21 | 92 ± 18 | 0.3 |
| ACE-I, n/total (%) | 0 | 0 | 0 | - |
| ARB, n/total (%) | 3/32 (9) | 1/8 (13) | 2/24 (8) | 1.0 |
| Diuretic, n/total (%) | 2/32 (6) | 0 | 2/24 (8) | 1.0 |
| Dose (mg), **median (IQR)** | 100 (IQR = 70-100) | 100 (IQR = 100-100) | 100 (IQR = 60-100) | 0.05 |
| Duration of Treatment (mo.), **median (IQR)** | 41 (IQR = 17-79) | 50 (IQR = 41-77) | 31 (IQR = 15-79) | 0.2 |

### Assessing Albuminuria on Dasatinib vs. other TKIs

Patients treated with dasatinib (n= 32) for at least 90 days had significantly higher UACR levels (median 28.0 mg/g, IQR 11.5 – 119.5) than patients treated with other TKIs (n=50; median 15.0 μg/mg, IQR 8.0 – 35.0; p = 0.03) for at least 90 days (**Figure 1**). Exactly 50% of all dasatinib-treated patients in this study exhibited at least moderately increased albuminuria, compared to only 30% for non-dasatinib treated CML patients; out of those who did develop albuminuria, 19% had severely increased albuminuria (UACR > 300 mg/ in the dasatinib cohort but 0% in the non-dasatinib group (**Figure 2**). There were no notable differences in baseline characteristics of those that developed albuminuria vs. those that did not while on dasatinib, other than duration of treatment with the TKI (50 months vs. 31 months). In those treated with dasatinib, the duration of exposure was significantly associated with albuminuria (ß= 1.2, SE= 0.4; *p* =0.003) (**Figure 3**).

**Figure 1.**
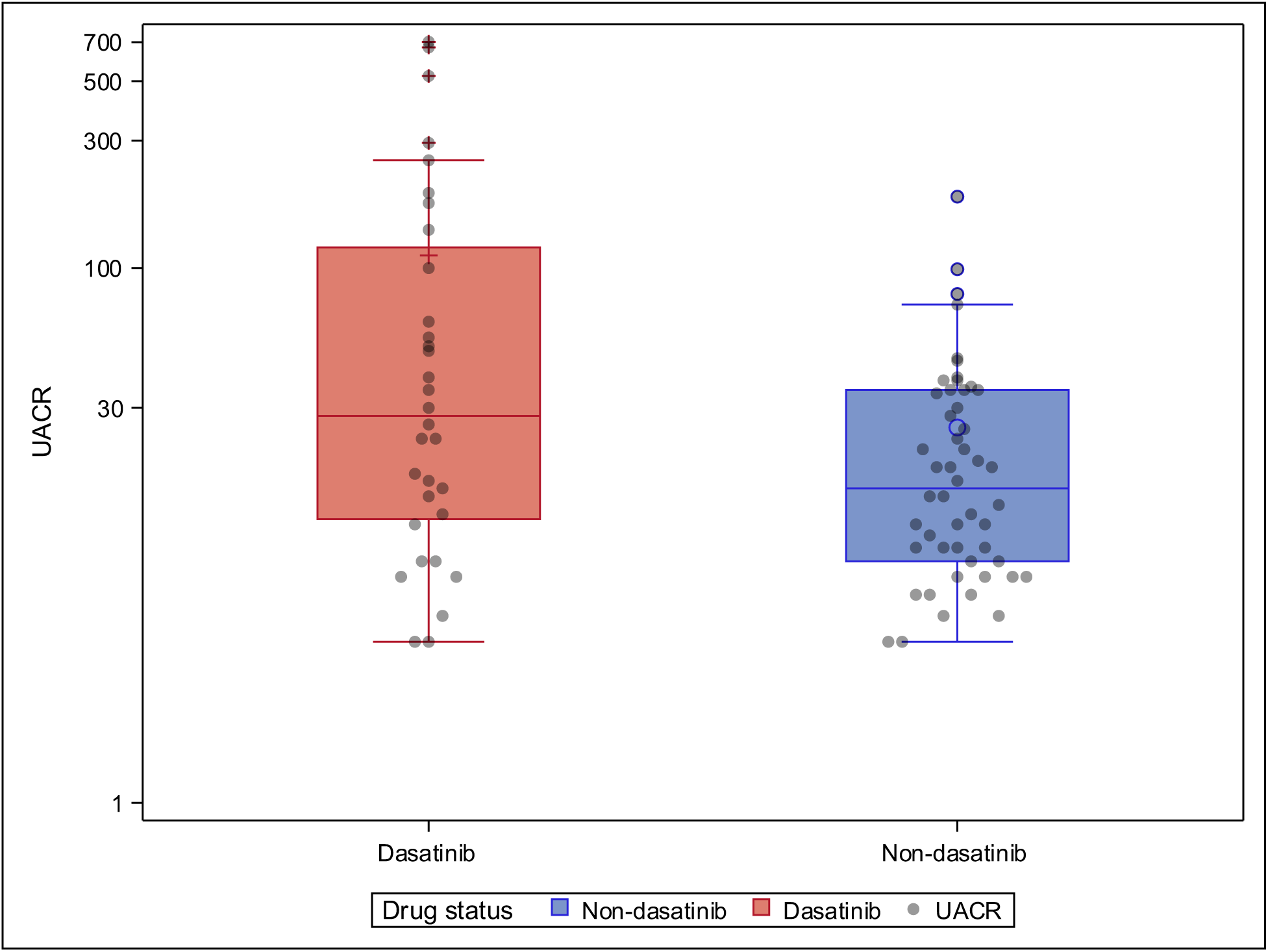
Boxplot for urine albumin-creatinine ratio (UACR) values for dasatinib and non-dasatinib patients. Chronic myeloid leukemia patients on dasatinib for at least 90 days (n= 32) had significantly higher UACR levels (median 28.0 μg/mg, IQR 11.5 – 119.5) than patients treated with other TKIs (imatinib, nilotinib, bosutinib and ponatinib) for at least 90 days (n=50; median 15.0 μg/mg, IQR 8.0 – 35.0; p = 0.03). Our case study patient had a UACR of 702.

**Figure 2.**
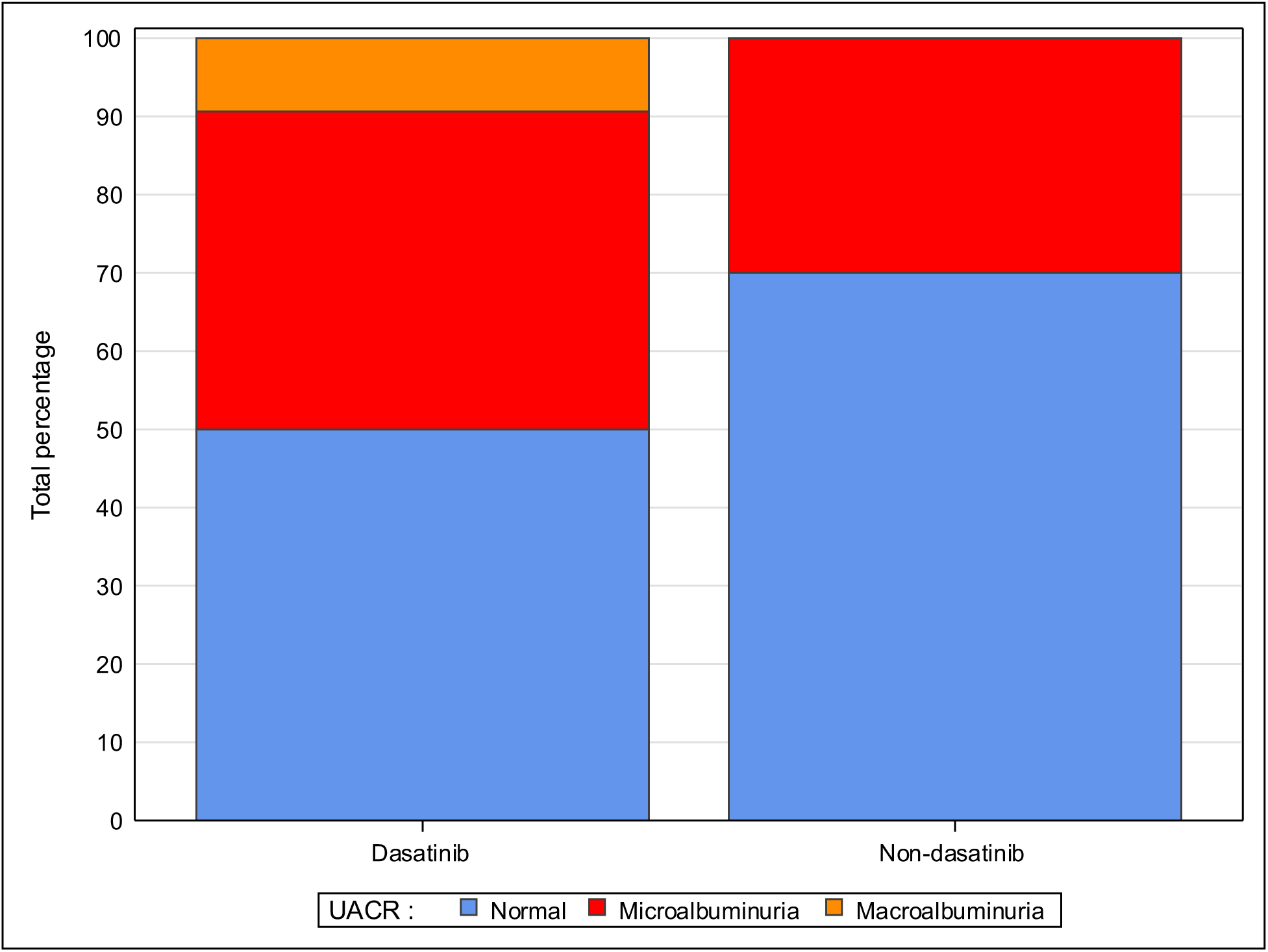
Percentages of patients with albuminuria in dasatinib and non-dasatinib patients. Half of all dasatinib treated patients in this study exhibited albuminuria, compared to 30% for non-dasatinib treated CML patients. 9% of dasatinib patients had sever albuminuria (> 300 mg/g), whereas none of the patients in the non-dasatinib group.

**Figure 3.**
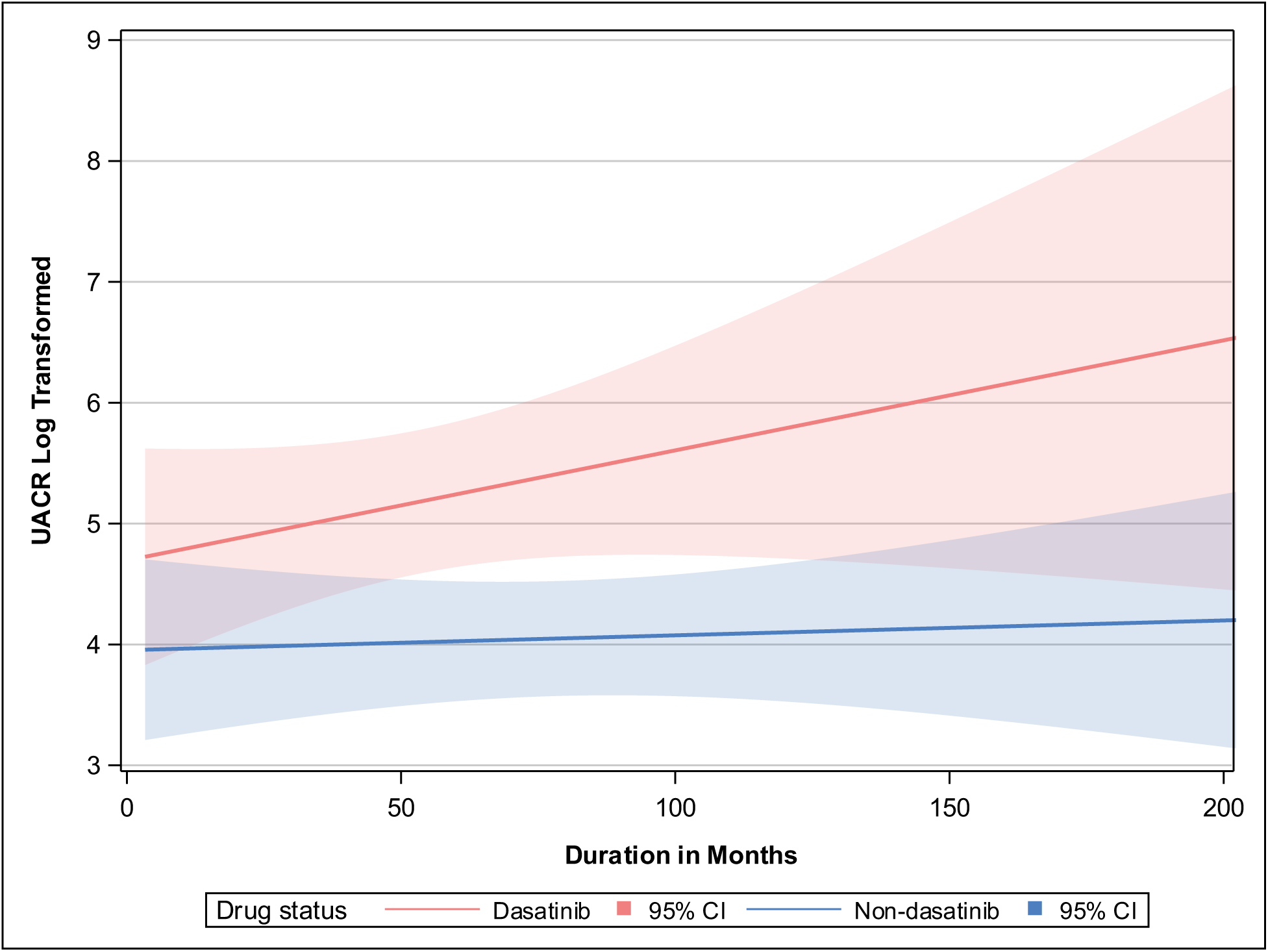
Level of UACR over treatment duration for dasatinib and non-dasatinib patients. The unadjusted estimates of log transformed UACR for patients being treated with dasatinib compared to non-dasatinib in interaction with time was insignificant. Estimate (SE) 0.01 (0.01), p-value = 0.3. However, trend of increased UACR as a function of treatment time in dasatinib patients is apparent.

### Pharmacokinetics

For the sixteen patients receiving dasatinib that had plasma obtained for analysis of drug concentration at the time of urine collection, dasatinib plasma concentration had a significant positive correlation with albuminuria (*r* = 0.54, p = 0.03; **Figure 4**). All other correlations are presented on Table 3.

**Table 3.** Spearman correlation coefficients (and p-values) among UACR and pharmocokinetic factors.

|  |  |  |  |  |  |
| --- | --- | --- | --- | --- | --- |
| **UACR** | 1.00 |  |  |  |  |
| **Dasatinib Plasma Concentration** | 0.54 (0.03) | 1.00 |  |  |  |
| **Dasatinib Oral Clearance** | -0.26 (0.33) | -0.60 (0.01) | 1.00 |  |  |
| **Dasatinib Max Steady State Concentration** | -0.02 (0.95) | 0.41 (0.12) | -0.48 (0.06) | 1.00 |  |
| **Dasatinib Average Steady State Concentration** | 0.17 (0.53) | 0.71 (0.002) | -0.86 (<0.001) | 0.7 (0.003) | 1.00 |
| **N=16** | **UACR** | **Dasatinib Plasma Concentration** | **Dasatinib Oral Clearance** | **Dasatinib Max Steady State Concentration** | **Dasatinib Average Steady State Concentration** |

**Figure 4.**
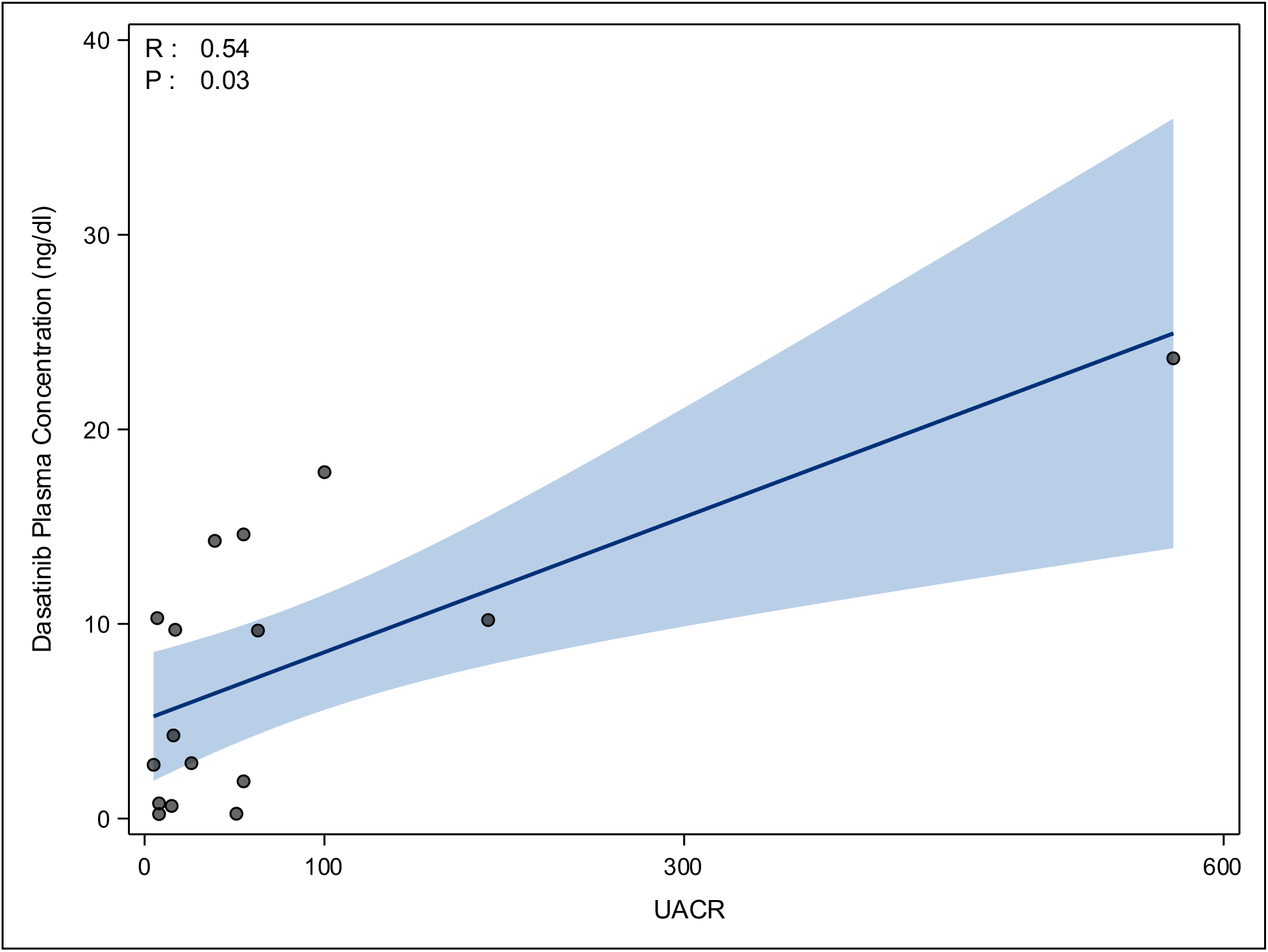
Significant and positive correlation was observed between dasatinib plasma concentration and UACR (R = 0.54, p-value = 0.03).

### Biopsy Results

One of the patients in the cohort underwent a kidney biopsy. She was a 43-year-old black female, who was diagnosed with CML in 2007. She had no known history of hypertension, heart failure, or diabetes. Initial management with imatinib began in October 2007 but was eventually discontinued in 2012 due to elevated liver function tests. She then began a course of nilotinib, which ended in 2013 after noting headaches and palpitations. Since then, she was started on 100 mg daily of dasatinib.

The biopsy participant’s urinalysis six years later showed a protein level above 1,000 mg/dL as well as a urine protein-creatinine (UPC) ratio of 3 (257/85). A follow-up urinalysis a week later was roughly unchanged, with a UPC ratio of 2.9 (470/159). She was admitted for a kidney biopsy about a month later, which showed chronic endothelial injury/chronic thrombotic microangiopathy (TMA) extensively involving glomeruli, global glomerulosclerosis, mild interstitial fibrosis and mild tubular atrophy (**Figures 5A and 5B**). Immunofluorescence showed no significant glomerular or tubulointerstitial staining for immunoglobulins, complement components, or kappa or lambda light chains. Lastly, electron microscopy showed widespread podocyte effacement of more than 80% of the basement membrane surfaces (**Figures 6A and 6B**).

**Figure 5.**
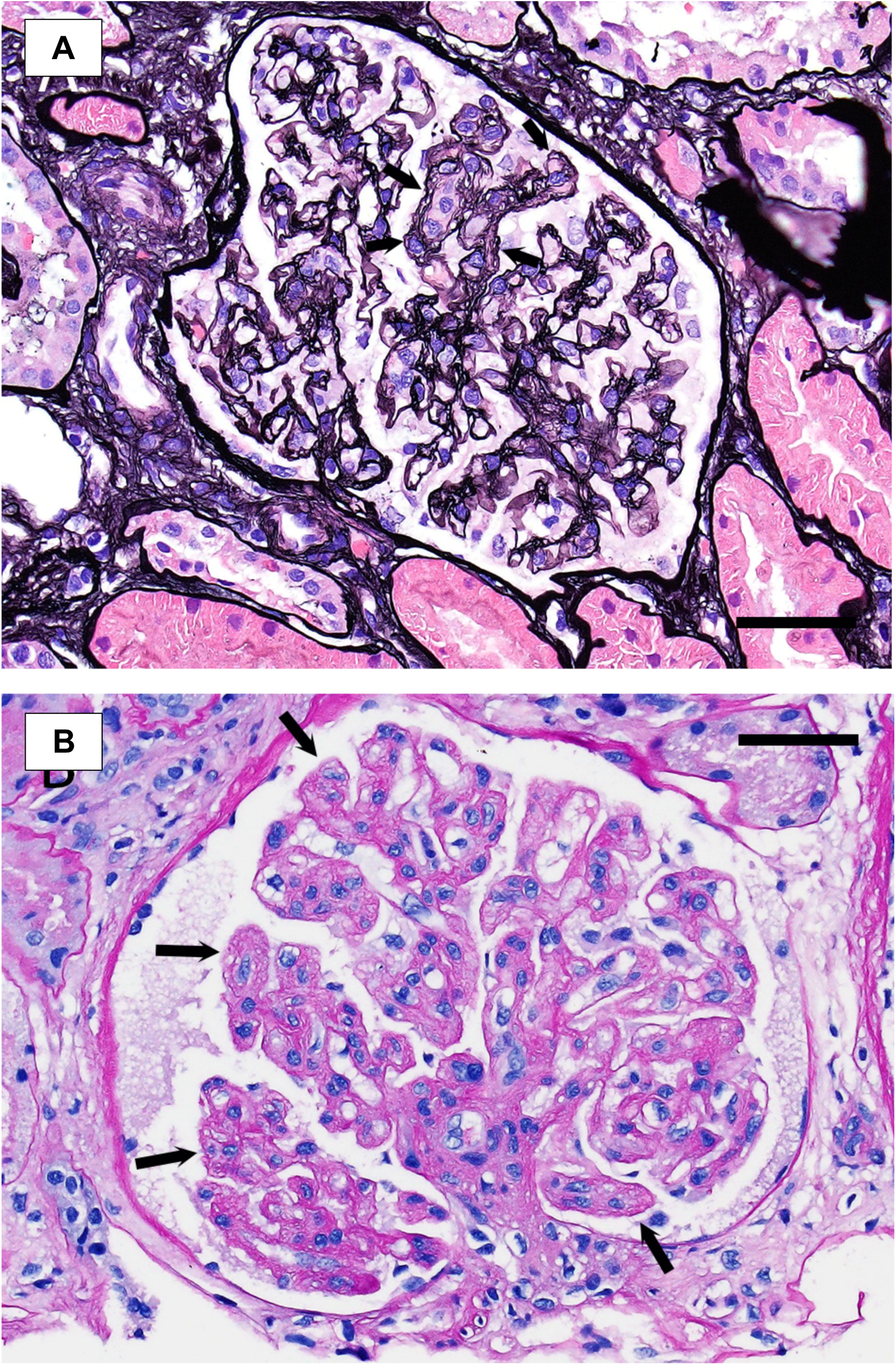
Glomerulus from dasatinib-treated biopsy patient showing segmental capillary wall double contour formation (arrows) in both **(A)** Jones Silver staining, 40x, as well as **(B)** PAS staining, 40x. Scale bars represent 50 μm.

**Figures 6.**
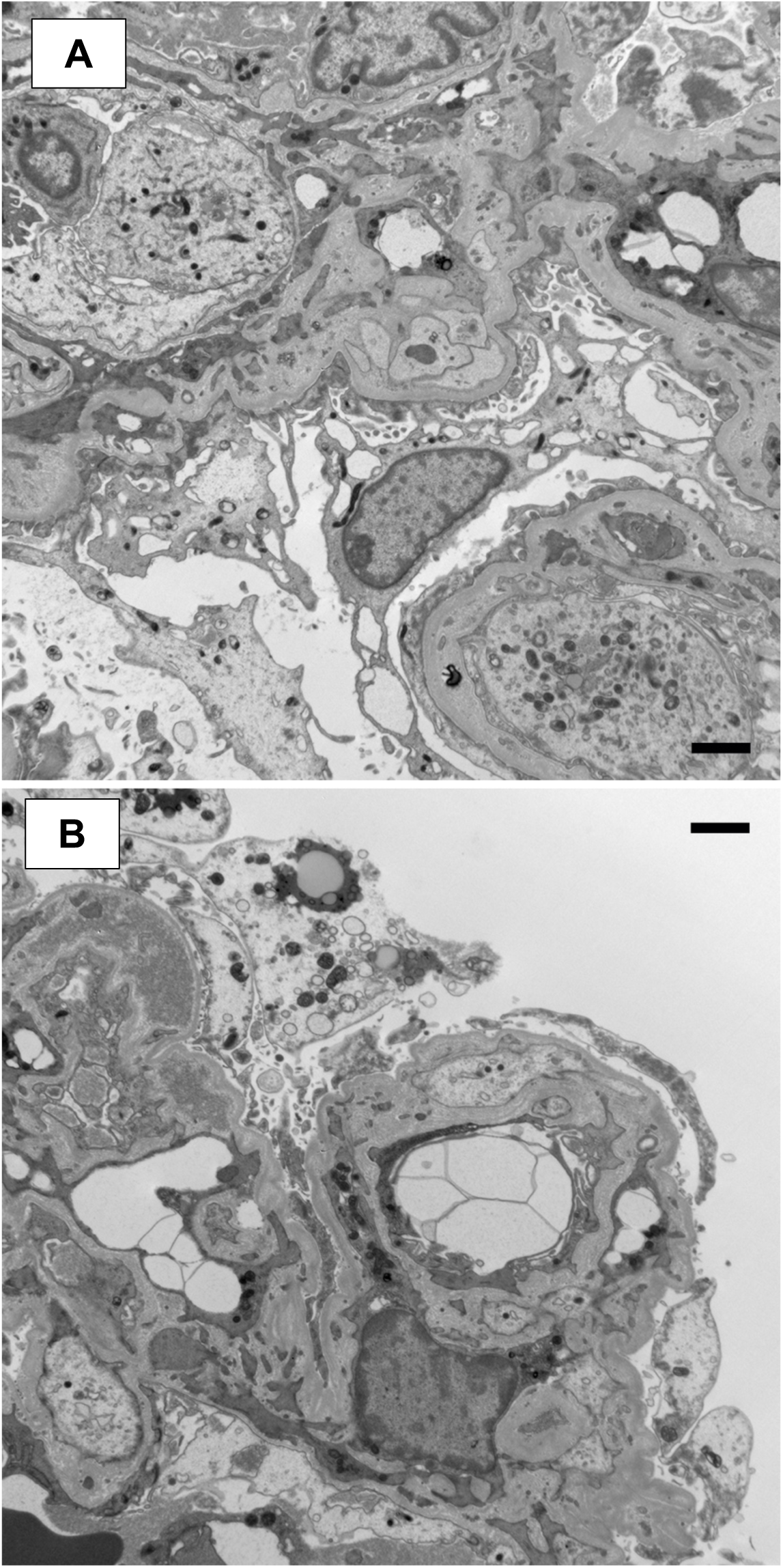
The glomeruli from a dasatinib-treated biopsy patient show global foot process effacement along with glomerular basement membrane duplication and multilayering along with focal entrapment of hyaline material but no deposits. Focal intracapillary mononuclear inflammatory cells can be seen **(A)** Transmission electronc microscopy, 6000×. **(B)** Transmission electron microscopy, 6000×. Scale bars represent 2 μm.

Based on these findings dasatinib was discontinued. Her follow-up UPC ratio about a week later was markedly decreased to 0.761G (115/151). Since then, she has been treated with 5 mg daily of lisinopril and had a repeat UPC ratio of 0.304G (28/92).

## Discussion

We found that CML patients receiving dasatinib were much more likely to have proteinuria compared to other TKI therapies. Despite being a younger and healthier cohort, half of all dasatinib treated patients in this study exhibited at least moderately increased albuminuria, compared to only 30% for non-dasatinib treated CML patients; out of those who did develop albuminuria, about 1 in 5 had severely increased albuminuria within the dasatinib cohort compared to none in the non-dasatinib TKI group. Glomerular damage documented in one case supports the concept that the glomerular changes caused by dasatinib are not only diffuse, but also reversible, as shown by the significant improvement in renal function after dasatinib discontinuation. The reversible nature of dasatinib-associated kidney injury is consistent with our original hypothesis of dasatinib-associated disruption of the podocyte actin cytoskeleton, which is reversible. It also emphasizes the need for vigilant monitoring for signs of proteinuria in patients receiving dasatinib since early intervention has the potential to avoid chronic irreversible kidney dysfunction.

Interestingly, non-dasatinib users had a statistically significant higher serum creatinine concentration and lower eGFR compared to dasatinib treated patients, which is likely due to non-dasatinib treated patients, on average, being approximately a decade older. Kidney function has been shown to naturally decrease with age ^15^. Non-dasatinib patients also had higher frequency of comorbidities. However, despite these factors for increased susceptibility of developing albuminuria, non-dasatinib patients still had a much lower incidence of mild albuminuria than their healthier dasatinib counterparts. This observation also supports the idea of dasatinib’s potent nephrotoxicity compared to its sister TKIs that have comparable kinase target profiles ^16,17^.

In addition, pharmacokinetic analyses showed that dasatinib metabolism and glomerular function may, in fact, be linked. Average steady state dasatinib concentration or maximum steady state concentration correlated moderately and significantly with UACR. These results might be explained by the fact that nephrotoxic dasatinib users received higher daily dasatinib doses than their non-nephrotoxic counterparts. As the first multicenter study to highlight potential renal toxicity of dasatinib on human glomeruli, our results showed the high prevalence of dasatinib related nephrotoxicity that supported our earlier laboratory findings ^8^. Importantly, cardiovascular and renal comorbidities were largely absent within the group of patients with proteinuria, which decreased the likelihood of secondary causes of proteinuria confounding the results. Taken together, all these findings strongly support our hypothesis that dasatinib directly affects glomerular podocytes.

In addition to the results from our study, the follow-up results from a dasatinib-treated biopsy patient further corroborates our hypothesis on dasatinib toxicity on the glomeruli. After the glomerular changes from the biopsy were discovered, dasatinib was discontinued for the patient. Interestingly, her follow-up UPC ratio about a week after discontinuation was markedly decreased from 2.9 to 0.761 (115/151). Since then, she has been treated with 5 mg daily of lisinopril, and she had a repeat UPC ratio of 0.304 (28/92). Her proteinuria did not completely disappear, however, as we believe that she has pre-existing chronic TMA most likely due to myeloproliferative neoplasm-induced glomerulopathy. Myeloproliferative neoplasm-induced glomerulopathy is chronic endothelial injury in the setting of long-standing clonal hematopoietic stem cell disorders, such as CML, polycythemia vera (PV), and essential thrombocythemia (ET) ^18^. Given that the biopsy patient had CML for over 10 years, it is very likely that she was predisposed to glomerular injury. However, the diffuse foot process effacement on EM and marked decrease of the UPC after dasatinib discontinuation suggests a secondary process related to dasatinib treatment, rather than her chronic TMA. Our biopsy case is compelling in that it not only is consistent with the results from our earlier mechanistic study with murine glomeruli, but it also stresses the need for careful monitoring of dasatinib treatment since there may also be patients with pre-existing chronic TMA, who will continue to unknowingly injure their kidney ^8^.

There are several limitations to our study, sample size being the first one. Although there are enough data to confirm increased proteinuria among dasatinib treated patients, we were unable to draw major conclusions with strong statistical significance with the current sample size. For example, our data does indeed show that the frequency of albuminuria in patients receiving dasatinib was greater than that of patients not receiving dasatinib. The pharmacokinetic analyses, which were incorporated later in the study timeline, showed several significant relationships; however, there were several inconclusive outcomes, also due to low sample size. Our smaller sample size was partly due to the self-imposed limitation of our cohort to CML patients as opposed to including other forms of Ph+ leukemia, such as ALL. Another limitation of our study is participant diversity. While the ages were fairly distributed, nearly 80% of the study patients were white. The lack of African American patients, for example, made it impossible to look at the link between *APOL1* and susceptibility to dasatinib nephrotoxicity, which would have been helpful to address its association as an additional risk factor ^19^. We also lacked repeated serial collections and assessment of UACR in the participants to analyze trends over time of exposure. Lastly, since this was a retrospective clinical study and not a place-controlled clinical trial, some patients had a history of multiple TKIs. We do not think this was a major constraint though, as the long duration of treatment for most patients would help mitigate contraindicative effects of earlier treatments.

In conclusion, dasatinib use in CML patients is shown to have a significantly higher likelihood of developing glomerular damage compared to non-dasatinib TKI treatments. Our results demonstrate that half of patients receiving dasatinib are at risk for developing at least moderately increased albuminuria, with 10% experiencing severely increased albuminuria. These findings warrant further prospective evaluation and updated guidelines for monitoring for this treatment emergent AE as well as preemptive approaches to mitigate the risk and potential consequences. Preemptive approaches may include screening guidelines, such as checking for UACR every month after initiation. Dasatinib is an effective therapeutic option for the treatment of CML; however, monitoring for the potential emergence of kidney glomerular damage in the form of proteinuria is warranted and should be studied in a prospective cohort.

## Data Availability

All data produced in the present study are available upon reasonable request to the authors.

## Acknowledgements

The study was supported by R01DK1182222 from the NIH (EUA) as well as PR202312 from the DoD, and R01DK129299 and R01GM123330 from the NIH (EAJ). Biospecimens and data from the Mount Sinai Hospital were provided through the Hematological Malignancies Tissue Bank (HMTB), an independent tissue bank repository for hematological malignancies and disorders with increased risk of developing into overt hematological malignancies. The HMTB is administered under the auspices of the NCI-designated Tisch Cancer Institute (TCI) at the Icahn School of Medicine at Mount Sinai.

## References

1. Cortes JE, Saglio G, Kantarjian HM, et al. Final 5-Year Study Results of DASISION: The Dasatinib Versus Imatinib Study in Treatment-Naïve Chronic Myeloid Leukemia Patients Trial. Journal of Clinical Oncology. 2016;34(20):2333–2340.

2. Cortes JE, Jiang Q, Wang J, et al. Dasatinib vs. imatinib in patients with chronic myeloid leukemia in chronic phase (CML-CP) who have not achieved an optimal response to 3 months of imatinib therapy: the DASCERN randomized study. Leukemia. 2020;34(8):2064–2073.

3. Jhaveri KD, Wanchoo R, Sakhiya V, Ross DW, Fishbane S. Adverse Renal Effects of Novel Molecular Oncologic Targeted Therapies: A Narrative Review. Kidney Int Rep. 2017;2(1):108–123.

4. Demetri GD, Lo Russo P, MacPherson IRJ, et al. Phase I Dose-Escalation and Pharmacokinetic Study of Dasatinib in Patients with Advanced Solid Tumors. Clinical Cancer Research. 2009;15(19):6232–6240.

5. Downing NS, Shah ND, Aminawung JA, et al. Postmarket Safety Events Among Novel Therapeutics Approved by the US Food and Drug Administration Between 2001 and 2010. JAMA. 2017;317(18):1854–1863.

6. Coca SG, Singanamala S, Parikh CR. Chronic kidney disease after acute kidney injury: a systematic review and meta-analysis. Kidney Int. 2012;81(5):442–448.

7. Piscitani L, Sirolli V, Di Liberato L, Morroni M, Bonomini M. Nephrotoxicity Associated with Novel Anticancer Agents (Aflibercept, Dasatinib, Nivolumab): Case Series and Nephrological Considerations. International Journal of Molecular Sciences. 2020;21(14).

8. Calizo RC, Bhattacharya S, van Hasselt JGC, et al. Disruption of podocyte cytoskeletal biomechanics by dasatinib leads to nephrotoxicity. Nat Commun. 2019;10(1):2061.

9. Zhao S, Nishimura T, Chen Y, et al. Systems pharmacology of adverse event mitigation by drug combinations. Science translational medicine. 2013;5(206):206ra140.

10. Ozkurt S, Temiz G, Acikalin MF, Soydan M. Acute renal failure under dasatinib therapy. Ren Fail. 2010;32(1):147–149.

11. Wallace E, Lyndon W, Chumley P, Jaimes EA, Fatima H. Dasatinib-induced nephrotic-range proteinuria. American journal of kidney diseases : the official journal of the National Kidney Foundation. 2013;61(6):1026–1031.

12. Ochiai S, Sato Y, Minakawa A, Fukuda A, Fujimoto S. Dasatinib-induced nephrotic syndrome in a patient with chronic myelogenous leukemia: a case report. BMC nephrology. 2019;20(1):87.

13. Ruebner RL, Copelovitch L, Evageliou NF, Denburg MR, Belasco JB, Kaplan BS. Nephrotic syndrome associated with tyrosine kinase inhibitors for pediatric malignancy: case series and review of the literature. Pediatr Nephrol. 2014;29(5):863–869.

14. Inker LA, Astor BC, Fox CH, et al. KDOQI US commentary on the 2012 KDIGO clinical practice guideline for the evaluation and management of CKD. American journal of kidney diseases : the official journal of the National Kidney Foundation. 2014;63(5):713–735.

15. Weinstein JR, Anderson S. The aging kidney: physiological changes. Adv Chronic Kidney Dis. 2010;17(4):302–307.

16. van Hasselt JGC, Rahman R, Hansen J, et al. Transcriptomic profiling of human cardiac cells predicts protein kinase inhibitor-associated cardiotoxicity. Nat Commun. 2020;11(1):4809.

17. Hantschel O, Grebien F, Superti-Furga G. The growing arsenal of ATP-competitive and allosteric inhibitors of BCR-ABL. Cancer Res. 2012;72(19):4890–4895.

18. Said SM, Leung N, Sethi S, et al. Myeloproliferative neoplasms cause glomerulopathy. Kidney Int. 2011;80(7):753–759.

19. Reidy KJ, Hjorten R, Parekh RS. Genetic risk of APOL1 and kidney disease in children and young adults of African ancestry. Curr Opin Pediatr. 2018;30(2):252–259.

